# Sex differences in the association of mild behavioral impairment with cognitive aging

**DOI:** 10.1101/2021.05.20.21257514

**Authors:** Katrin Wolfova, Byron Creese, Dag Aarsland, Zahinoor Ismail, Anne Corbett, Clive Ballard, Adam Hampshire, Pavla Cermakova

## Abstract

**INTRODUCTION:** We aimed to explore sex differences in the association of mild behavioral impairment (MBI) with the level of cognitive performance and its rate of decline in a cohort of people without dementia with the longest term follow up of cognition.

**METHODS:** We studied 8,181 older adults enrolled in the online PROTECT UK Study. MBI was assessed using the MBI Checklist and cognition was measured by digit span, paired associate learning, spatial working memory and verbal reasoning. Statistical analysis was conducted using linear regression models and linear mixed-effects models.

**RESULTS:** Males exhibited more often symptoms of decreased motivation, impulse dyscontrol and social inappropriateness, while less often symptoms of emotional dysregulation. The associations of MBI domains with some measures of cognitive performance and decline was stronger in males than females, with the exception of emotional dysregulation.

**DISCUSSION:** MBI may influence cognition to a greater extent in males than in females.

## 1. BACKGROUND

Females bear a higher burden of dementias than men in terms of prevalence, incidence, and also severity.^1^ Two thirds of patients with Alzheimer disease (AD)^2^, the most common cause of dementia, are females.^3^ While multiple cognitive abilities are affected by AD more adversely in females^4^, males experience steeper cognitive decline when affected by other forms of dementia.^5^ Sex differences in cognitive functioning exist also in healthy older adults, with males performing better in some visuospatial tasks, whereas females outperform males in tasks focused on verbal domains.^6^ Many new biomarker studies are rapidly emerging but only few consider sex as a moderator.^7^ It is crucial to understand the contribution of sex in the variations in cognitive aging to optimize prevention and intervention strategies.

There is a need to develop more sensitive measures to accurately identify individuals at a higher risk of incident cognitive decline. Mild behavioral impairment (MBI) is a promising indicator of a population at risk. MBI is characterized by a persistent change in personality or behavior starting later in life, represented by the emergence of neuropsychiatric symptoms (NPS) such as apathy, anxiety, mood disturbances, agitation, disinhibition, lack of empathy, loss of insight, or psychosis.^8^ MBI is associated with faster cognitive and functional decline.^9,10^ Also individual NPS, especially anxiety and irritability, have been found to be related to cognitive decline.^11,12^ Moreover, several studies report links between MBI and neurobiological markers, such as higher β-amyloid deposition, tau pathology, and neurodegeneration in pre-dementia population ^13-15^. While sex differences in non-cognitive manifestations are well described in dementia population,^16-18^ little is known about these differences in individuals without dementia.

Previous studies suggest that the prevalence of MBI – specifically decreased motivation, impulse dyscontrol, apathy, agitation, and irritability - in a cognitively healthy population might be higher in males^19-21^. It is plausible to hypothesize that sex might play a role not only in the prevalence of individual NPS but might also moderate the relationship between NPS and cognition in dementia-free populations. In the present study, we aimed to explore sex differences in MBI and its associations with the level of cognitive performance and the rate of cognitive decline in a large sample of community-dwelling dementia-free individuals from the United Kingdom.

## 2. METHODS

### 2.1. Source of data

We utilized data from the Platform for Research Online to Investigate Genetics and Cognition in Aging (PROTECT, https://www.protectstudy.org.uk/), a longitudinal online research project that collects data from ∼25,000 healthy individuals in the United Kingdom. The first wave of data collection started in November 2015 and has been followed by subsequent assessments on a yearly basis. In addition, refreshment samples are added annually to keep the sample size steady. Eligible participants 1) are based in the United Kingdom, 2) have a good understanding of English, 3) are 50 years old or over, 4) have regular access to a computer and the internet, and 5) were not diagnosed with dementia. All participants provided informed consent via online platform and all data were pseudo-anonymized. The study was approved by London Bridge National Research Ethics Committee (Reference: 13/LO/1578).

### 2.2. Mild behavioral impairment

MBI was assessed using the Mild Behavioral Impairment Checklist (MBI-C) rated by informants who have known the participant for at least 10 years. MBI-C comprises 34 questions developed to evaluate the presence and severity of NPS in healthy and pre-dementia populations.^22^ The symptom is considered present if it represents a change from longstanding behavior and persists for at least 6 months (continuously or intermittently). Individual domains of MBI are: 1) decreased motivation, decreased interest and drive, apathy (which will be further on referred as “decreased motivation”); 2) emotional or affective dysregulation, mood and anxiety symptoms (“emotional dysregulation”); 3) impulse dyscontrol, agitation, aggression, and abnormal reward salience (“impulse dyscontrol”); 4) social inappropriateness, impaired social cognition (“social inappropriateness”); and 5) abnormal thoughts and perception, psychotic symptoms (“psychotic symptoms”).

The severity of each present symptom is rated: 1 = mild (noticeable, but not a significant change); 2 = moderate (significant, but not a dramatic change); and 3 = severe (very marked or prominent, a dramatic change). We excluded 156 participants with incomplete data in any of the MBI domains (Supplemental Figure S1). We created a binary variable representing MBI syndrome using a cut-off value of more than 8 points (“MBI syndrome”), which has demonstrated good sensitivity and specificity for clinically diagnosed MBI according to the ISTAART diagnostic criteria in participants with subjective cognitive decline,^10^ and binary variables indicating the presence of at least one symptom of any severity in an individual MBI domain, which have also been used in previous studies.^23^

### 2.3. Cognition

Cognition was measured on a yearly basis using four tests that had been previously adapted and validated for online use: digit span (DS), paired associate learning (PAL), self-ordered search (SOS) and verbal reasoning (VR).^24,25^ The participants had up to three attempts to complete the same test battery across seven days ensuring a break of 24 hours between sessions. The score of DS, PAL and SOS represents the total number of correct answers. The score of VR test is calculated as the total number of trials answered correctly, minus the number answered incorrectly. The final outcome measures were calculated as a mean of the scores from up to the three separate attempts. Cognitive decline was operationalized as an annual decrease in the final scores by including a two-way interaction between time and exposure in longitudinal analysis.

The DS test is based on the ability to remember a sequence of numbers that appear on the screen one at a time and captures deficits in attention and concentration.^26^ The PAL test evaluates episodic memory by presenting a series of objects that are hidden under boxes. Participants are required to remember, which object is hidden under which box. Deficits in PAL have been found to correlate with hippocampal atrophy, which is one of the first neuroimaging features of AD.^27^ The SOS test is based on self-ordered search tasks for assessment of spatial working memory. The task is to find a symbol hidden under an on-screen box and, when found, participants are asked to search for another symbol while remembering that a symbol would not be hidden in the same box twice. Lower scores often indicate frontal lobe damage.^28^ The VR test is an online adaptation of Baddeley’s Gramatical Reasoning test, which correlates well with the ability to reason, analyse and solve problems known as fluid intelligence.^29^ Participants are asked to click on “True” or “False” button in order to express whether they agree with a statement describing a relationship between two shapes (a circle, a square, etc.) on the screen.

### 2.4. Covariates

Covariates were selected based on literature as sociodemographic and health-related characteristics associated with MBI and cognition.^30^ Sociodemographic characteristics include information on sex (male vs. female), ethnic origin (white vs. non-white), co-habitation status (married/co-habiting vs. living alone), employment status (employed vs. other). Education level was categorized into three groups: low (secondary education); middle (post-secondary education, vocational qualification, undergraduate degree) and high (post-graduate degree, doctorate). Socioeconomic covariates were categorized for the purpose of descriptive analysis, detailed description of covariates used in statistical models is included in the Supplement. Health-related characteristics were self-reported at baseline and include body-mass index (BMI; calculated as weight in kilograms divided by height in meters squared), hypertension, history of heart disease (heart disease, heart attack and/or angina), diabetes and hypercholesterolemia.

### 2.5. Analytical sample

Only participants that had 1) available measures on cognition at baseline and at least 1 follow-up occasion, and 2) informant rated MBI-C assessment at baseline or at year 1, if baseline assessment was not performed, were considered for the present analysis (n = 10,244). To assure that presence of MBI is not attributable to other health conditions, we excluded participants who reported history of stroke (n=149), Parkinson’s disease (n=22), mania/bipolar/manic depression (n=48), anxiety/generalized anxiety disorder (n=1 410), social anxiety (n=121), agoraphobia (n=34), panic attacks (n=520), obsessive-compulsive disorder (n=34), anorexia nervosa (n=81), bulimia nervosa (n=54), psychological overeating/binge eating (n=50), schizophrenia (n=5), other psychotic illnesses (n=28), personality disorders (n=13), autism/Asperger’s/autistic spectrum disorder (n=13), or attention deficit disorder (n=7). In addition, participants with clinical signs of depression defined by score ≥ 14 at Patient Health Questionnaire-9 scale were also excluded (n=99). This procedure is in accordance with the International Society to Advance Alzheimer’s Research and Treatment-Alzheimer’s Association (ISTAART-AA) MBI criteria and has been used in previous studies.^10,22^ In total, 2,063 participants were excluded (some participants reported more than one of the above-mentioned conditions), leaving 8,181 persons in the final analytical sample (flowchart presented on Supplemental Figure S1) with median follow-up 3.07 years (interquartile range 2.02-3.22). Participants with missing data on covariates were kept in the sample for the descriptive analyses.

### 2.6. Statistical analysis

The analysis was performed in two steps. First, we tested whether the association of MBI (syndrome and individual MBI domains) with the level of cognitive performance differs by sex (*cross-sectional analysis*). Second, we studied whether males and females differ in the association of MBI (syndrome and individual MBI domains) with the rate of cognitive decline (*longitudinal analysis*).

#### 2.6.1. Cross-sectional analysis

Descriptive data of the analytical sample is presented as frequency (n [%]), mean ± standard deviation (SD), or median and interquartile range (IQR). Differences between males and females were tested using independent samples t-test for continuous variables with normal distribution, Mann-Whitney test for continuous variables with skewed distribution and χ2 test for binary variables. Effect size of the sex difference was calculated as Cohen’s d for continuous variables and Cramer’s v for categorical variables. Linear regression was applied to estimate beta coefficients with 95% confidence intervals (CIs) for the associations of the independent variable MBI (syndrome and individual MBI domains) with the level of cognitive performance at baseline.

To assess whether sex moderates the association of MBI (syndrome and individual MBI domains) with the level of cognitive performance, we included a two-way interaction term between MBI (syndrome and individual MBI domains) and sex into a model adjusted for age and assessed the interaction effect using likelihood ratio (LR) test. We performed stratified analyses, where appropriate. Three sets of models are presented, stepwise adjusting for covariates. Model 1 is adjusted for age, Model 2 for age and sociodemographic characteristics (employment status, ethnic origin, co-habitation status, education level), Model 3 for age, sociodemographic and health-related characteristics (BMI, hypertension, history of heart disease, diabetes, hypercholesterolemia).

#### 2.6.2. Longitudinal analysis

Methods of linear mixed-effects modelling were applied to explore whether there is an association of MBI (syndrome and individual MBI domains) with the rate of cognitive decline. Participants and time (in years since baseline) were set as random intercepts, time as random slope at participant level, and time, MBI, sex and baseline age (centered around mean) as fixed effects. To assess whether sex moderates the association of MBI with the rate of cognitive decline, we included a three-way interaction term between MBI, time and sex (MBI × time × sex) in Model 1. We performed stratified analyses, where appropriate. We added other covariates as fixed effects in a stepwise approach: sociodemographic characteristics (employment status, ethnic origin, co-habitation status, education level) in Model 2; and health-related characteristics (BMI, hypertension, history of heart disease, diabetes, hypercholesterolemia) in Model 3. In addition, as participants might become familiar with the cognitive test battery when it is administered repeatedly, which could mask cognitive decline, we controlled for practice effect. To select the right method to control for practice effect, we compared three sets of models using different indicators in Model 1: 1) binary indicator of the first test; 2) number of prior tests; and 3) the square root of the number of prior tests.^31^ The indicator based on square root of the number of prior tests performed best according to Akaike information criterion and is therefore used in all presented models.

#### 2.6.3. Secondary analysis

We performed two sets of secondary analyses. First, because the prevalence of NPS is lower in younger individuals with AD^32,33^, we assumed the occurrence of MBI might follow the same age group pattern. In addition, recent evidence shows that there is a stronger association between genetic markers and cognitive decline in older age groups.^34^ To assess whether there are age differences in the association of MBI with cognitive performance, we tested a three-way interaction of MBI and sex with age group (55-65 vs. 65 and more) and then we stratified the analysis by age groups and sex, if appropriate. Second, because people with mild cognitive impairment exhibit symptoms of MBI more often than cognitively healthy individuals^35^, we repeated the longitudinal analysis on a dataset of a cognitively healthy population (n=7,853 participants). We excluded participants who self-reported mild cognitive impairment (n=15) at baseline and participants with a baseline level of cognitive performance 1.5 or more standard deviations (indicating mild cognitive impairment below the average on 2 or more cognitive domains (n=325).

As this was an exploratory analysis, we did not control for multiple comparisons and use p value <0.05 as a threshold for statistical significance. All analyses were carried out using Stata software v 16.1.

## 3. RESULTS

Out of 8 181 individuals (median age 63 years, 73% females), 11% of females and 14% of males had a score of more than 8 points on MBI-C (p=0.014, V=0.036, Table 1). Females and males differed in the proportion of 4 out of 5 individual MBI domains: females exhibited more often symptoms of emotional dysregulation (45% vs. 36% in males; p<0.001), while less often symptoms of decreased motivation (25% vs. 30%; p<0.001), impulse dyscontrol (40% vs. 44%; p=0.001) and social inappropriateness (12% vs. 15%; p<0.001). There were no significant sex differences in frequency of psychotic symptoms. There were sex differences with a small effect size in the scores of all 4 cognitive tests: females scored lower than males in DS (7.41 vs. 7.54; p=0.001, d=-0.085), PAL (4.54 vs. 4.50; p=0.03, d=0.052) and SOS (7.54 vs. 7.88; p<0.001, d=-0.158), but higher in VR (32.69 vs. 31.87; p<0.001, d=0.093).

**Table 1.** Sex differences in baseline characteristics.

|  | **Females**<br>**(n=5,970, 73.0%)** | **Males**<br>**(n= 2,211, 27.0%)** | **p value<sup>a</sup>** | **Effect size<sup>b</sup>** |
| --- | --- | --- | --- | --- |
| Age, median (IQR) | 61.8 (56.9–66.8) | 64.8 (59.5–69.5) | <0.001 | -0.370 |
| White ethnic origin, n (%) | 5 569 (93.3) | 2 089 (94.5) | 0.041 | -0.022 |
| Married/co-habiting, n (%) | 4 603 (77.1) | 1 979 (89.6) | <0.001 | -0.139 |
| Education level, n (%) |  |  | <0.001 | 0.054 |
| Low | 755 (12.65) | 286 (12.9) |  |  |
| Middle | 3 963 (66.4) | 1 354 (61.3) |  |  |
| High | 1 252 (21.0) | 570 (25.8) |  |  |
| Employed, n (%) | 2 506 (42.0) | 799 (36.2) | <0.001 | 0.053 |
| Cognition |  |  |  |  |
| Digit span, mean ± SD | 7.41 ± 1.48 | 7.54 ± 1.45 | 0.001 | -0.085 |
| Paired associate learning, mean ± SD | 4.54 ± 0.76 | 4.50 ± 0.74 | 0.031 | 0.052 |
| Verbal reasoning, mean ± SD | 32.70 ± 8.98 | 31.87 ± 8.70 | <0.001 | 0.093 |
| Spatial working memory, mean ± SD | 7.55 ± 2.04 | 7.88 ± 2.26 | <0.001 | -0.158 |
| MBI |  |  |  |  |
| MBI syndrome (< 8 points), n (%) | 656 (10.99) | 300 (13.57) | 0.001 | 0.036 |
| Decreased motivation, n (%) | 1 508 (25.26) | 669 (30.26) | <0.001 | 0.050 |
| Emotional dysregulation, n (%) | 2 701 (44.8) | 797 (35.6) | <0.001 | -0.082 |
| Impulse dyscontrol, n (%) | 2 350 (39.36) | 962 (43.51) | 0.001 | 0.038 |
| Social inappropriateness, n (%) | 704 (11.79) | 336 (15.20) | <0.001 | 0.045 |
| Psychotic symptoms, n (%) | 405 (6.78) | 127 (5.74) | 0.090 | -0.018 |
| BMI, median (IQR) | 24.2 (22.1–27.3) | 25.3 (23.2–27.8) | <0.001 | -0.152 |
| Comorbidities |  |  |  |  |
| Hypertension, n (%) | 1 420 (23.85) | 741 (33.71) | <0.001 | 0.099 |
| Heart disease, n (%) | 160 (2.7) | 184 (8.4) | <0.001 | 0.126 |
| Diabetes, n (%) | 135 (2.3) | 117 (5.3) | <0.001 | 0.078 |
| Hypercholesterolemia, n (%) | 211 (3.5) | 122 (5.6) | <0.001 | 0.045 |

SD, standard deviation; IQR, interquartile range; MBI, mild behavioral impairment; BMI, body-mass index a t-test for continuous variables with normal distribution, Mann-Whitney-Wilcoxon test for continuous variables with skewed distribution or chi-square test for categorical variables
b effect size of the sex difference is presented as Cohen's d for continuous variables and Cramer's v for categorical variables

### 3.1. Cross-sectional analysis

We found several associations of MBI with the level of baseline cognitive performance across individual domains in the whole analytical sample (Supplemental Table S1). Sex moderated the association of the MBI syndrome with the level of cognitive performance measured by PAL (p from LR test 0.035). The MBI syndrome was associated with a lower level of PAL score only in males (B -0.158; 95% CI -0.245 to -0.072; Model 1, Table 2). The association attenuated but remained statistically significant in the fully adjusted model (B -0.154; 95% CI -0.241 to -0.067; Model 3, Table 2). When considering individual MBI domains, sex moderated the association of impulse dyscontrol with the level of cognitive performance measured by DS (p from LR test 0.040) and by PAL (p from LR test 0.035). When stratified, impulse dyscontrol was associated with a lower level of DS score only in males (B=-0.229; 95% CI -0.351 to - 0.108, Model 1, Table 2) and with PAL score only in males (B=-0.093; 95% CI -0.153 to - 0.033, Model 1, Table 2). All associations attenuated but remained significant after adjustment for other covariates.

**Table 2.** Association of mild behavioral impairment with the level of cognitive performance, stratified by sex.

**B (95% CI)**
|  | Digit span |  | Paired associate learning |  |
| --- | --- | --- | --- | --- |
|  | Females | Males | Females | Males |
| MBI syndrome |  |  |  |  |
| Model 1 | - | - | -0.036 (-0.096;<br>0.024) | -0.158 (-0.245;<br>-0.072)*** |
| Model 2 | - | - | -0.029 (-0.089;<br>0.032) | -0.153 (-0.240;<br>-0.066)*** |
| Model 3 | - | - | -0.023 (-0.084;<br>0.038) | -0.154 (-0.241;<br>-0.067)*** |
| Impulse dyscontrol |  |  |  |  |
| Model 1 | -0.074 (-0.150;<br>0.003) | -0.229 (-0.351;<br>-0.108)*** | -0.019 (-0.058;<br>0.020) | -0.093 (-0.153;<br>-0.033)** |
| Model 2 | -0.066 (-0.143;<br>0.011) | -0.200 (-0.321;<br>-0.078)** | -0.016 (-0.055;<br>0.023) | -0.080 (-0.141;<br>-0.020)** |
| Model 3 | -0.061 (-0.138;<br>0.016) | -0.200 (-0.321;<br>-0.080)** | -0.014 (-0.053;<br>0.024) | -0.080 (-0.140;<br>-0.019)** |

\*\*\* p<0.001 \*\* p<0.01 \* p<0.05; CI, confidence interval; MBI, mild behavioral impairment
Model 1: baseline age
Model 2: baseline age, employment status, ethnic origin, co-habitation status, education level
Model 3: baseline age, employment status, ethnic origin, co-habitation status, education level, body-mass index, hypertension, history of heart disease, diabetes, hypercholesterolemia

### 3.2. Longitudinal analysis

We found an association of the MBI syndrome and all MBI domains with cognitive decline measured by SOS and VR (Supplemental Table S3) in the whole analytical sample. Sex moderated the association of MBI syndrome as well as all MBI domains with the rate of cognitive decline measured by VR (p from LR test <0.001). When stratified by sex, the MBI syndrome (Supplemental Figure S2), decreased motivation and impulse dyscontrol were related to a higher rate of cognitive decline in VR in both sexes, but in all cases to a greater extent in males than females (Table 3). The association of emotional dysregulation with the rate of cognitive decline in VR was present exclusively in females (B=-0.175; 95% CI -0.297 to -0.052, Model 1, Table 3), while the association of social inappropriateness (B=-0.298; 95% CI -0.564 to -0.031, Model 1, Table 3) and psychotic symptoms (B=-0.554; 95% CI -0.977 to -0.132, Model 1, Table 3) with a higher rate of cognitive decline was present exclusively in males. These associations attenuated but remained statistically significant in the fully adjusted model.

**Table 3.** Association of mild behavioral impairment with the rate of decline in verbal reasoning, stratified by sex.

|  | B (95% CI) |  |
| --- | --- | --- |
|  | Verbal reasoning |  |
|  | Females | Males |
| MBI syndrome |  |  |
| Model 1 | -0.282 (-0.479; -0.084)** | -0.324 (-0.604; -0.045)* |
| Model 2 | -0.282 (-0.479; -0.084)** | -0.325 (-0.604; -0.046)* |
| Model 3 | -0.273 (-0.472; -0.074) ** | -0.323 (-0.602; -0.043)* |
| Emotional dysregulation |  |  |
| Model 1 | -0.175 (-0.297; -0.052)** | -0.178 (-0.376; 0.021) |
| Model 2 | -0.175 (-0.297; -0.052)** | -0.176 (-0.375; 0.022) |
| Model 3 | -0.172 (-0.295; -0.049)** | -0.177 (-0.376; 0.022) |
| Decreased motivation |  |  |
| Model 1 | -0.229 (-0.371; -0.088)** | -0.334 (-0.541; -0.127)** |
| Model 2 | -0.231 (-0.372; -0.090)** | -0.334 (-0.541; -0.128)** |
| Model 3 | -0.223 (-0.364; -0.081)** | -0.335 (-0.542; -0.128)** |
| Impulse dyscontrol |  |  |
| Model 1 | -0.135 (-0.260; -0.010)* | -0.216 (-0.407; -0.024)* |
| Model 2 | -0.135 (-0.260; -0.011)* | -0.216 (-0.407; -0.024)* |
| Model 3 | -0.133 (-0.259; -0.008)* | -0.211 (-0.403; -0.019)* |
| Social inappropriateness |  |  |
| Model 1 | -0.188 (-0.378; 0.003) | -0.298 (-0.564; -0.031)* |
| Model 2 | -0.190 (-0.380; 0.001) | -0.297 (-0.563; -0.031)* |
| Model 3 | -0.186 (-0.378; 0.005) | -0.283 (-0.550; -0.016)* |
| Psychotic symptoms |  |  |
| Model 1 | -0.160 (-0.408; 0.087) | -0.554 (-0.977; -0.132)* |
| Model 2 | -0.159 (-0.406; 0.088) | -0.557 (-0.978; -0.135)** |
| Model 3 | -0.143 (-0.392; 0.106) | -0.553 (-0.976; -0.131)* |

\*\*\* p<0.001 \*\* p<0.01 \* p<0.05; CI, confidence interval; MBI, mild behavioral impairment
Model 1: baseline age, practice effect
Model 2: baseline age, practice effect, employment status, ethnic origin, co-habitation status, education level
Model 3: baseline age, practice effect, employment status, ethnic origin, co-habitation status, education level, body-mass index, hypertension, history of heart disease, diabetes, hypercholesterolemia

### 3.3. Secondary analysis

There was a significant interaction (p value from LR test < 0.05) with age group in cognitive performance measured by PAL. When stratified by age group, the associations found in the cross-sectional analysis were present only in the group of older males (65 years or older, Supplemental Table S2). When the longitudinal analysis was repeated on a sample of cognitively healthy individuals, we obtained results similar to our main findings: significant associations between higher rate of cognitive decline in VR and MBI domains were present only in males (Supplemental Table S4 and S5). Specifically, we found these associations in the domain of decreased motivation and psychotic symptoms in all three models.

## 4. DISCUSSION

In the present study, we explored sex differences in the association of MBI with cognitive ageing for the first time. Overall, males exhibited symptoms in more domains of MBI than females, particularly decreased motivation, impulse dyscontrol and social inappropriateness. Total MBI-C score and individual domains were associated with cognitive performance and rate of decline in the whole sample. However, when sex differences were present, these associations were present either exclusively in males or to a greater extent in males, with the exception of emotional dysregulation, which was present only in females.

Our results add valuable insights into connections between sex, MBI, and cognition. Mortby et al. reported that males were more likely to exhibit symptoms of decreased motivation and impulse dyscontrol than females.^19^ Similarly, apathy, agitation and irritability have been reported more frequently in older males than females in other studies.^20,21^ Given that dementia is a syndrome caused by several distinctive underlying diseases affecting different brain areas, our novel focus on the putative prodromal marker of MBI results suggest that non-cognitive symptoms of dementia may vary even during early manifestation.

A possible explanation of our findings might lie in the entorhinal cortex. A study based on a sample from the Baltimore Longitudinal Study of Aging shows that males with amyloid pathology, a marker of AD, experience a steeper volumetric decline in entorhinal and parahippocampal regions, thus suggesting that females are more resilient towards volumetric loss in these areas.^36^ This hypothesis is supported by recent findings from the Czech Brain Aging Study, which revealed that atrophy in both the entorhinal cortex and hippocampus is associated with MBI in a group of memory clinic patients with subjective cognitive decline and mild cognitive impairment.^37^ In addition, higher tau-PET signal in the entorhinal cortex/hippocampus is associated with higher MBI-C scores in cognitively unimpaired amyloid positive individuals.^14^

Another explanation could be the fact that individual dementia subtypes presenting with different NPS are not equally distributed between sexes. For example, psychotic symptoms affect around 30% of patients with AD in comparison to around 50% of patients with PD dementia and dementia with Lewy bodies (DLB), both of which are more common in males than females.^38-40^ Recently, late onset psychiatric symptoms have been proposed as one of the three prototypic prodromal DLB syndromes.^41^ In addition, decreased motivation and psychotic symptoms have been found to be strong predictors of disease progression in frontotemporal degeneration (FTD), which appears to be more common in males.^42-44^ Moreover, recent findings show that females with behavioral variant of FTD have greater ability to cope with neuropathological changes compared to males and they exhibit less behavioral symptoms, including apathy, despite the same level of atrophy burden.^45^ In contrast, progression of AD, in which females account for 60% of patients^2^, has been more often linked to the presence of depressive symptoms.^44,46^

Several limitations need to be mentioned. Participants were not representative of the general population of the United Kingdom, as females, individuals with higher education and White people were overrepresented. Next, we performed a large number of statistical tests, which increases the risk of type I error, but most of our comparisons are likely to be correlated. Furthermore, the observed effect sizes are relatively small, but meaningful on a population level. On the contrary, this study is unique as MBI has not been used as a predictor of cognitive decline separately for males and females before. We used a battery of four neuropsychological tests assessing multiple cognitive domains to find all possible links between MBI and cognition. Even though the follow-up period for a cognitively healthy community dwelling sample is rather short in our study, to our knowledge, it represents the longest and most detailed neuropsychological analysis of MBI to date. Further longitudinal studies with biomarker assessment are required to find out the role of sex in the relationship between MBI symptoms and dementia pathology.

In conclusion, this study provides unique evidence that there are sex differences in the prevalence of MBI symptoms as well as in the association of MBI with the level of cognitive performance and its rate of decline in older adults. Males who exhibit MBI symptoms are at risk of faster cognitive decline than females, particularly when psychotic symptoms and symptoms of decreased motivation occur.

## Data Availability

The data that support the findings of this study are available from but restrictions apply to the availability of these data, which were used under license for the current study, and so are not publicly available. Data are however available from the authors upon reasonable request and with permission of the PROTECT study.

## ACKNOWLEDGMENTS

This paper represents independent research coordinated by the University of Exeter and King’s College London and is funded in part by the National Institute for Health Research (NIHR) Biomedical Research Centre at South London and Maudsley NHS Foundation Trust and King’s College London. This research was also supported by the National Institute for Health Research (NIHR) Collaboration for Leadership in Applied Health Research and Care South West Peninsula and the National Institute for Health Research (NIHR) Exeter Clinical Research Facility. The views expressed are those of the authors and not necessarily those of the NHS, the NIHR or the Department of Health.

The authors were supported by the Ministry of Health of the Czech Republic (Grant NU20J-04-00022; PC, KW), PRIMUS (247066) conducted at Charles University (PC, KW), the “Sustainability for the National Institute of Mental Health” (Grant LO1611), with a financial support from the Ministry of Education, Youth and Sports of the Czech Republic (PC), and by the Czech Alzheimer Endowment Fund (KW).

