## Supplement for "Sex differences in the association of mild behavioral impairment with cognitive aging"

**Description of covariates**

Ethnic origin

Participants were asked “What is your ethnic origin?” and were offered the following options: White: English / Welsh / Scottish / Northern Irish / British, White: Irish; White: Gypsy or Irish Traveller; White: European; White: Non-European; Mixed: White and Black Caribbean; Mixed: White and Black African; Mixed: White and Asian; Mixed: Any other Mixed / Multiple ethnic background; Asian / Asian British: Indian; Asian / Asian British: Pakistani; Asian / Asian British: Bangladeshi; Asian / Asian British: Chinese; Asian / Asian British: Any other Asian background; Black / African / Caribbean / Black British: African; Black / African / Caribbean / Black British: Caribbean; Any other Black / African / Caribbean background; Other ethnic group: Arab; Any other ethnic group.

Employment status

Participants were asked “What is your current employment status?” and were offered the following options: Employed (full time); Employed (part time); Self-employed, Retired; Unemployed.

Marital status

Participants were asked “What is your marital status?” and were offered the following options: Married; Widowed; Separated; Divorced; Civil Partnership; Co-habiting; Single.

Education

Participants were asked “What is the highest level of education you have completed?” and were offered the following options: Secondary Education (GCSE/O-Levels); Post-Secondary Education (College, A-Levels, NVQ3 or below, or similar); Vocational Qualification (Diploma, Certificate, BTEC, NVQ 4 and above, or similar); Undergraduate Degree (BA, BSc etc.); Post-graduate Degree (MA, MSc etc.); Doctorate (PhD).

BMI

Participants were asked “What is your height?” and “What is your current weight?”. BMI was calculated as weight in kilograms divided by height in meters squared.

Hypertension

Participants were asked “Has a doctor ever given you a diagnosis of, or told you that you have, any of the following?:” and were offered to answer “High blood pressure” as one of the options.

Heart disease

Participants were asked “Has a doctor ever given you a diagnosis of, or told you that you have, any of the following?:” and were offered to answer “Heart disease / Heart attack / Angina” as one of the options.

Diabetes

Participants were asked “Has a doctor ever given you a diagnosis of, or told you that you have, any of the following?:” and were offered to answer “Diabetes” as one of the options.

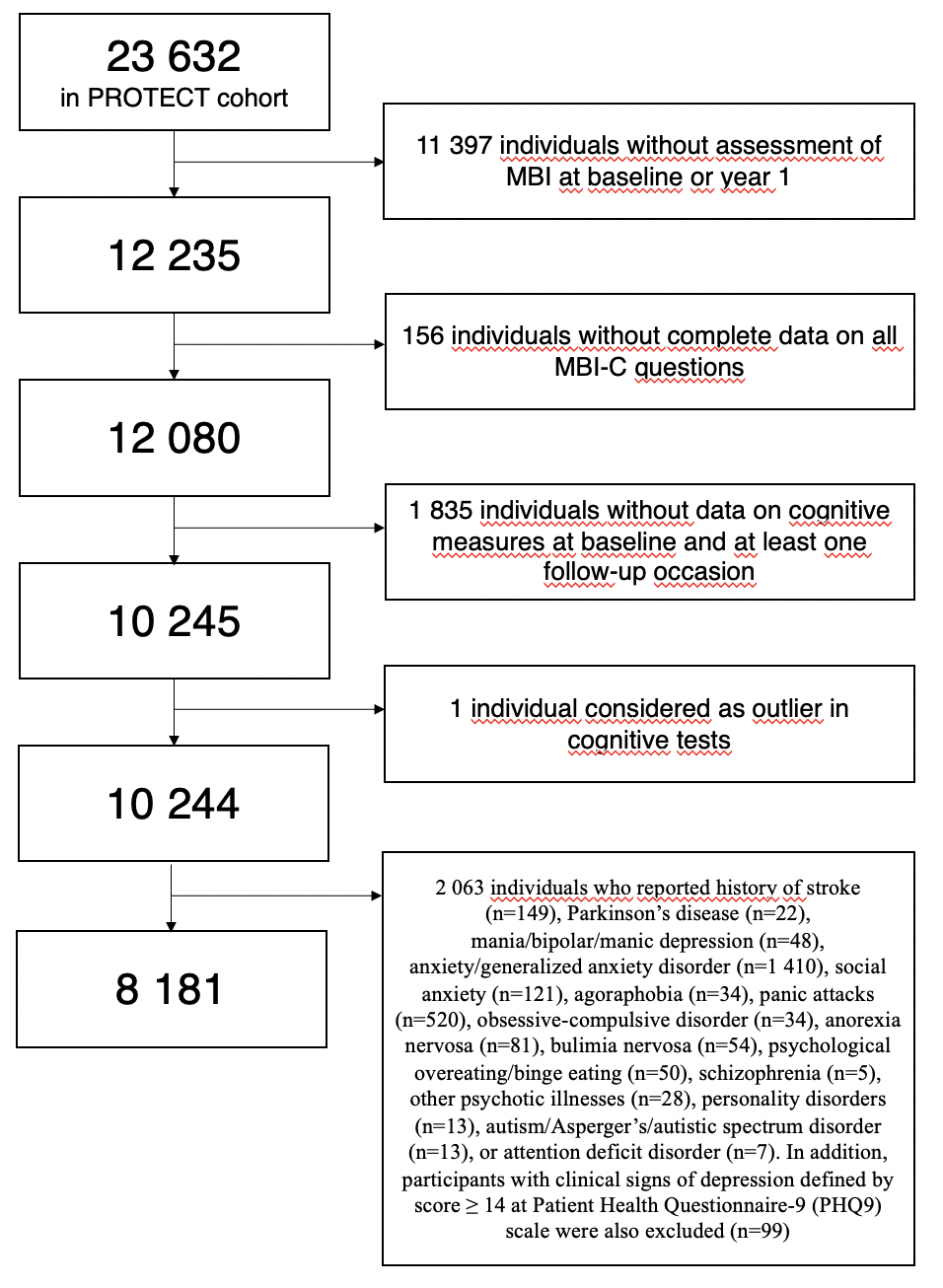

**Supplemental Figure S1** Selection of participants

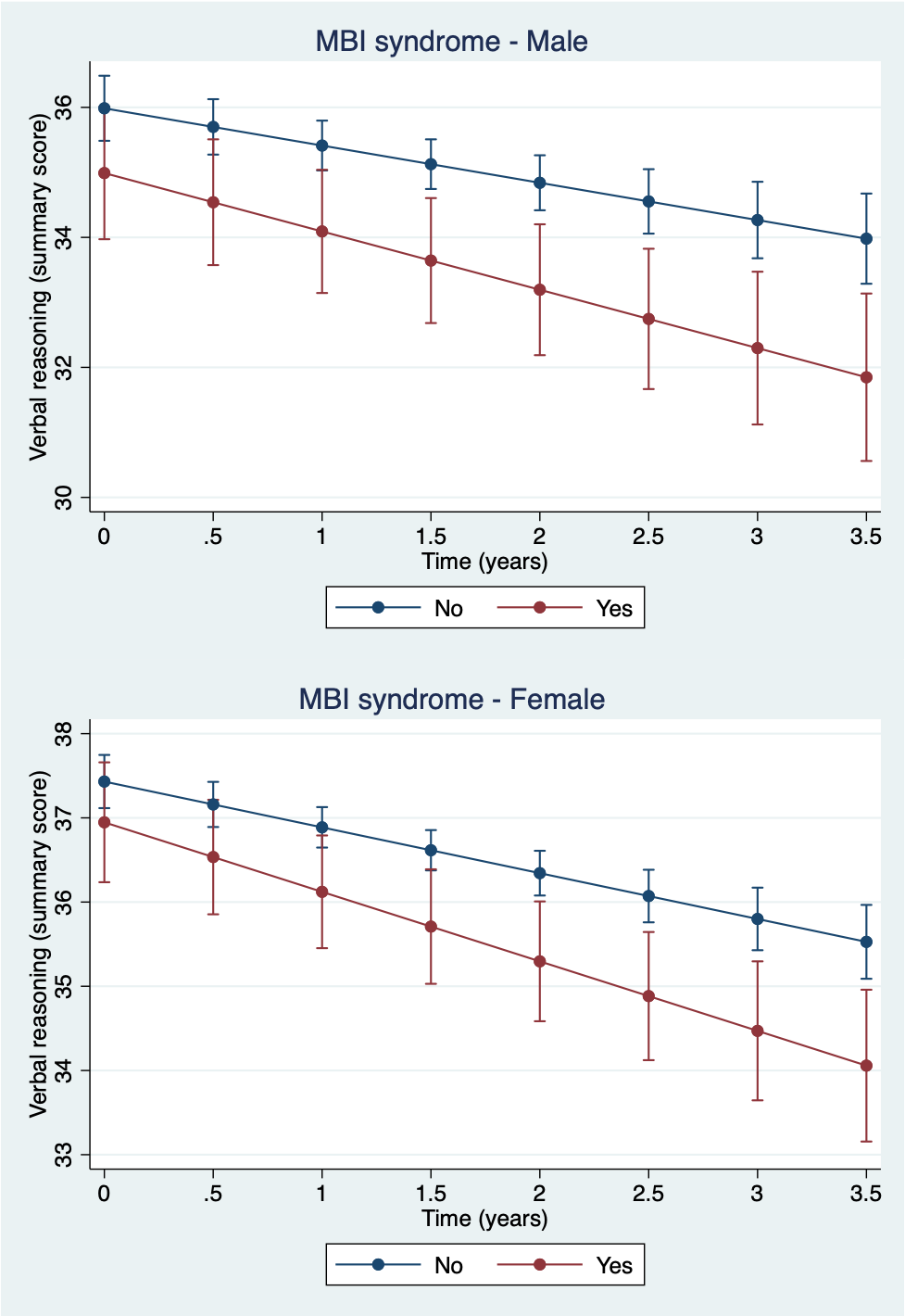

**Supplemental Figure S2** Predictive margins of MBI stratified by sex, Model 1

| **Supplemental Table S1** Association of MBI with cognitive performance in the whole analytical sample | | | | |
| --- | --- | --- | --- | --- |
|  | **B (95% CI)** | | | |
|  | **Digit span** | **Paired associate learning** | **Self-ordered search** | **Verbal reasoning** |
| MBI syndrome |  |  |  |  |
| Model 1 | -0.100 (-0.199;  -0.001)* | -0.074 (-0.124;  -0.025)** | -0.163 (-0.302;  -0.023)* | -0.508 (-1.088; 0.073) |
| Model 2 | -0.078 (-0.177; 0.021) | -0.068 (-0.117;  -0.018)** | -0.138 (-0.278; 0.001) | -0.197 (-0.762; 0.367) |
| Model 3 | -0.082 (-0.181; 0.018) | -0.064 (-0.114;  -0.014)* | -0.125 (-0.265; 0.015) | -0.160 (-0.729; 0.410) |
| Emotional dysregulation |  |  |  |  |
| Model 1 | -0.150 (-0.215;  -0.086)*** | -0.055 (-0.087;  -0.023)*** | -0.163 (-0.254;  -0.072)*** | -0.257 (-0.636; 0.122) |
| Model 2 | -0.146 (-0.211;  -0.082)*** | -0.054 (-0.087;  -0.022)*** | -0.151 (-0.242;  -0.060)** | -0.205 (-0.574; 0.164) |
| Model 3 | -0.140 (-0.205;  -0.076)*** | -0.051 (-0.084;  -0.019)** | -0.146 (-0.237;  -0.054)** | -0.197 (-0.567; 0.174) |
| Decreased motivation |  |  |  |  |
| Model 1 | -0.156 (-0.228;  -0.084)*** | -0.021 (-0.057; 0.015) | -0.077 (-0.179; 0.024) | -0.035 (-0.387; 0.458) |
| Model 2 | -0.140 (-0.212;  -0.068)*** | -0.015 (-0.051; 0.021) | -0.060 (-0.161; 0.042) | 0.227 (-0.184; 0.639) |
| Model 3 | -0.137 (-0.209;  -0.065)*** | -0.014 (-0.050; 0.022) | -0.043 (-0.145; 0.059) | 0.234 (-0.181; 0.648) |
| Impulse dyscontrol |  |  |  |  |
| Model 1 | -0.116 (-0.180;  -0.051)*** | -0.040 (-0.073;  -0.008)* | -0.112 (-0.204;  -0.021)* | -0.437 (-0.817;  -0.057)* |
| Model 2 | -0.102 (-0.167;  -0.037)** | -0.036 (-0.068;  -0.003)* | -0.098 (-0.189;  -0.007)* | -0.228 (-0.598; 0.142) |
| Model 3 | -0.100 (-0.165;  -0.035)** | -0.034 (-0.067;  -0.001)* | -0.091 (-0.183;  0.001) | -0.219 (-0.590; 0.153) |
| Social inappropriateness |  |  |  |  |
| Model 1 | -0.082 (-0.177; 0.014) | -0.043 (-0.091; 0.005) | -0.085 (-0.220; 0.049) | -0.628 (-1.189;  -0.067)* |
| Model 2 | -0.066 (-0.162; 0.029) | -0.037 (-0.085; 0.011) | -0.065 (-0.200; 0.070) | -0.408 (-0.954; 0.138) |
| Model 3 | -0.070 (-0.165; 0.026) | -0.040 (-0.088; 0.009) | -0.074 (-0.210; 0.061) | -0.427 (-0.976; 0.121) |
| Psychotic symptoms |  |  |  |  |
| Model 1 | -0.157 (-0.286;  -0.028)* | -0.080 (-0.144;  -0.015)* | -0.181 (-0.363;  0.000) | -0.880 (-1.636;  -0.124)* |
| Model 2 | -0.133 (-0.262;  -0.004)* | -0.067 (-0.132;  -0.003)* | -0.138 (-0.320;  0.044) | -0.537 (-1.273; 0.200) |
| Model 3 | -0.129 (-0.258; 0.000) | -0.064 (-0.129;  -0.001) | -0.140 (-0.322; -0.043) | -0.563 (-1.303; 0.178) |

*** p<0.001 ** p<0.01 * p<0.05; B, beta; CI, confidence interval; MBI, mild behavioral impairment

Model 1: baseline age, sex

Model 2: baseline age, sex, employment status, ethnic origin, co-habitation status, education level

Model 3: baseline age, sex, employment status, ethnic origin, co-habitation status, education level, body-mass index, hypertension, history of heart disease, diabetes, hypercholesterolemia

| **Supplemental Table S2** Association of mild behavioral impairment with the level of cognitive performance, stratified by sex and age group | | | |
| --- | --- | --- | --- |
|  | **B (95% CI)** | | |
|  | **Paired associate learning** | | |
|  | **Females** | | **Males** |
| **55-64** |  | |  |
| MBI syndrome |  |  | |
| Model 1 | -0.076 (-0.150; -0.003)* | -0.086 (-0.207; 0.035) | |
| Model 2 | -0.067 (-0.141; 0.007) | -0.079 (-0.202; 0.045) | |
| Model 3 | -0.058 (-0.133; 0.017) | -0.086 (-0.211; 0.038) | |
| Impulse dyscontrol |  |  | |
| Model 1 | -0.044 (-0.092; 0.003) | -0.059 (-0.143; 0.025) | |
| Model 2 | -0.040 (-0.087; 0.008) | -0.054 (-0.139; 0.030) | |
| Model 3 | -0.038 (-0.085; 0.010) | -0.054 (-0.139; 0.032) | |
| **≥ 65** |  |  | |
| MBI syndrome |  |  | |
| Model 1 | 0.051 (-0.054; 0.155) | -0.232 (-0.355; -0.108)*** | |
| Model 2 | 0.060 (-0.045; 0.165) | -0.228 (-0.352; -0.104)*** | |
| Model 3 | 0.059 (-0.048; 0.165) | -0.220 (-0.345; -0.096)*** | |
| Impulse dyscontrol |  |  | |
| Model 1 | 0.035 (-0.032; 0.101) | -0.134 (-0.220; -0.048)** | |
| Model 2 | 0.040 (-0.026; 0.107) | -0.119 (-0.206; -0.032)** | |
| Model 3 | 0.042 (-0.025; 0.109) | -0.118 (-0.206; -0.031)** | |
| *** p<0.001 ** p<0.01 * p<0.05; CI, confidence interval; MBI, mild behavioral impairment  Model 1: baseline age  Model 2: baseline age, employment status, ethnic origin, co-habitation status, education level  Model 3: baseline age, employment status, ethnic origin, co-habitation status, education level, body-mass index, hypertension, history of heart disease, diabetes, hypercholesterolemia | | | |

| **Supplemental Table S3** Association of MBI with the rate of cognitive decline in the whole analytical sample | | | | |
| --- | --- | --- | --- | --- |
|  | **B (95% CI)** | | | |
|  | **Digit span** | **Paired associate learning** | **Self-ordered search** | **Verbal reasoning** |
| MBI syndrome |  |  |  |  |
| Model 1 | -0.028 (-0.056; 0.000)* | 0.001 (-0.020; 0.021) | -0.079 (-0.131;  -0.027)** | -0.318 (-0.480;  -0.155)*** |
| Model 2 | -0.028 (-0.056; 0.000)* | 0.001 (-0.020; 0.021) | -0.078 (-0.130;  -0.027)** | -0.318 (-0.480;  -0.156)*** |
| Model 3 | -0.027 (-0.055; 0.001) | -0.004 (-0.024; 0.017) | -0.079 (-0.131;  -0.027)** | -0.313 (-0.477;  -0.150)*** |
| Emotional dysregulation |  |  |  |  |
| Model 1 | 0.000 (-0.018; 0.018) | -0.004 (-0.017; 0.009) | -0.049 (-0.082;  -0.016)** | -0.145 (-0.250;  -0.041)** |
| Model 2 | 0.001 (-0.017; 0.019) | -0.004 (-0.017; 0.009) | -0.048 (-0.082;  -0.015)** | -0.145 (-0.249;  -0.040)** |
| Model 3 | 0.001 (-0.017; 0.019) | -0.005 (-0.018; 0.008) | -0.048 (-0.081;  -0.015)** | -0.143 (-0.248;  -0.038)** |
| Decreased motivation |  |  |  |  |
| Model 1 | 0.002 (-0.018; 0.022) | -0.007 (-0.022; 0.008) | -0.068 (-0.105;  -0.030)*** | -0.283 (-0.400;  -0.166)*** |
| Model 2 | 0.002 (-0.018; 0.022) | -0.007 (-0.022; 0.008) | -0.068 (-0.106;  -0.031)*** | -0.284 (-0.401;  -0.167)*** |
| Model 3 | 0.001 (-0.019; 0.021) | -0.009 (-0.024; 0.006) | -0.071 (-0.108;  -0.033)*** | -0.280 (-0.398;  -0.163)*** |
| Impulse dyscontrol |  |  |  |  |
| Model 1 | 0.004 (-0.014; 0.022) | -0.006 (-0.020; 0.007) | -0.064 (-0.098;  -0.031)*** | -0.174 (-0.279;  -0.069)** |
| Model 2 | 0.004 (-0.014; 0.022) | -0.006 (-0.020; 0.007) | -0.064 (-0.098;  -0.030) *** | -0.174 (-0.279;  -0.069)** |
| Model 3 | 0.004 (-0.014; 0.022) | -0.007 (-0.021; 0.006) | -0.063 (-0.096;  -0.029)*** | -0.171 (-0.277;  -0.066)** |
| Social inappropriateness |  |  |  |  |
| Model 1 | 0.000 (-0.027; 0.026) | -0.019 (-0.039; 0.001) | -0.066 (-0.116;  -0.017)** | -0.249 (-0.405;  -0.092)** |
| Model 2 | -0.001 (-0.028; 0.026) | -0.019 (-0.039; 0.001) | -0.067 (-0.117;  -0.017)** | -0.250 (-0.406;  -0.094)** |
| Model 3 | 0.003 (-0.024; 0.029) | -0.018 (-0.037; 0.002) | -0.063 (-0.113;  -0.013)* | -0.243 (-0.400;  -0.086)** |
| Psychotic symptoms |  |  |  |  |
| Model 1 | 0.008 (-0.029; 0.045) | -0.000 (-0.027; 0.027) | 0.021 (-0.048; 0.090) | -0.236 (-0.451;  -0.022)* |
| Model 2 | 0.008 (-0.029; 0.045) | -0.000 (-0.028; 0.027) | 0.020 (-0.048; 0.089) | -0.236 (-0.450;  -0.022)* |
| Model 3 | 0.008 (-0.029; 0.045) | -0.002 (-0.029; 0.025) | 0.024 (-0.045; 0.093) | -0.225 (-0.440;  -0.010)* |

*** p<0.001 ** p<0.01 * p<0.05; B, beta; CI, confidence interval; MBI, mild behavioral impairment

Model 1: baseline age, sex, practice effect

Model 2: baseline age, sex, practice effect, employment status, ethnic origin, co-habitation status, education level

Model 3: baseline age, sex, practice effect, employment status, ethnic origin, co-habitation status, education level, body-mass index, hypertension, history of heart disease, diabetes, hypercholesterolemia

| **Supplemental Table S4** Association of MBI with the rate of cognitive decline in cognitively healthy analytical sample | | | | |
| --- | --- | --- | --- | --- |
|  | **B (95% CI)** | | | |
|  | **Digit span** | **Paired associate learning** | **Self-ordered search** | **Verbal reasoning** |
| MBI syndrome |  |  |  |  |
| Model 1 | -0.029 (-0.058; -0.001) * | 0.003 (-0.018; 0.024) | -0.083 (-0.136;  -0.030)** | -0.315 (-0.481;  -0.150) *** |
| Model 2 | -0.029 (-0.058; -0.001) * | 0.003 (-0.018; 0.024) | -0.083 (-0.135;  -0.030)** | -0.316 (-0.481;  -0.151) *** |
| Model 3 | -0.028 (-0.056; 0.000) | -0.002 (-0.022; 0.019) | -0.083 (-0.136;  -0.030)** | -0.311 (-0.477;  -0.145) *** |
| Emotional dysregulation |  |  |  |  |
| Model 1 | -0.003 (-0.021; 0.015) | -0.005 (-0.018; 0.009) | -0.052 (-0.086;  -0.019)** | -0.143 (-0.249;  -0.037)** |
| Model 2 | -0.003 (-0.021; 0.015) | -0.004 (-0.018; 0.009) | -0.052 (-0.085;  -0.018)** | -0.143 (-0.249;  -0.037)** |
| Model 3 | -0.002 (-0.020; 0.016) | -0.006 (-0.019; 0.008) | -0.051 (-0.085;  -0.017)** | -0.140 (-0.247;  -0.034)** |
| Decreased motivation |  |  |  |  |
| Model 1 | 0.001 (-0.020; 0.021) | -0.005 (-0.020; 0.010) | -0.063 (-0.101;  -0.025)** | -0.278 (-0.397;  -0.159)*** |
| Model 2 | 0.001 (-0.020; 0.021) | -0.005 (-0.020; 0.010) | -0.064 (-0.101;  -0.026)*** | -0.279 (-0.398;  -0.160)*** |
| Model 3 | 0.000 (-0.020; 0.021) | -0.007 (-0.022; 0.008) | -0.066 (-0.103;  -0.028)*** | -0.275 (-0.394;  -0.155)*** |
| Impulse dyscontrol |  |  |  |  |
| Model 1 | 0.000 (-0.019; 0.018) | -0.008 (-0.022; 0.005) | -0.070 (-0.104;  -0.036)*** | -0.166 (-0.273;  -0.059)** |
| Model 2 | 0.000 (-0.018; 0.018) | -0.008 (-0.022; 0.005) | -0.070 (-0.103;  -0.036)*** | -0.166 (-0.272;  -0.059)** |
| Model 3 | 0.000 (-0.019; 0.018) | -0.009 (-0.023; 0.004) | -0.069 (-0.102;  -0.035)*** | -0.163 (-0.270;  -0.056)** |
| Social inappropriateness |  |  |  |  |
| Model 1 | -0.002 (-0.029; 0.025) | -0.016 (-0.036; 0.004) | -0.063 (-0.113;  -0.012)* | -0.265 (-0.423;  -0.107)** |
| Model 2 | -0.002 (-0.029; 0.025) | -0.016 (-0.036; 0.004) | -0.063 (-0.113;  -0.013)* | -0.265 (-0.423;  -0.107)*** |
| Model 3 | 0.001 (-0.026; 0.028) | -0.014 (-0.034; 0.006) | -0.059 (-0.109;  -0.009)* | -0.259 (-0.417;  -0.100)** |
| Psychotic symptoms |  |  |  |  |
| Model 1 | 0.003 (-0.035; 0.040) | -0.007 (-0.035; 0.020) | 0.029 (-0.040; 0.099) | -0.220 (-0.438;  -0.001)* |
| Model 2 | 0.003 (-0.035; 0.040) | -0.007 (-0.035; 0.020) | 0.028 (-0.041; 0.098) | -0.219 (-0.437;  -0.001)* |
| Model 3 | 0.003 (-0.035; 0.040) | -0.010 (-0.037; 0.018) | 0.032 (-0.038; 0.102) | -0.207 (-0.427; 0.012) |

*** p<0.001 ** p<0.01 * p<0.05; B, beta; CI, confidence interval; MBI, mild behavioral impairment

Model 1: baseline age, sex, practice effect

Model 2: baseline age, sex, practice effect, employment status, ethnic origin, co-habitation status, education level

Model 3: baseline age, sex, practice effect, employment status, ethnic origin, co-habitation status, education level, body-mass index, hypertension, history of heart disease, diabetes, hypercholesterolemia

| **Supplemental Table S5** Association of MBI with the rate of decline in verbal reasoning in cognitively healthy analytical sample, stratified by sex | | |
| --- | --- | --- |
|  | **B (95% CI)** | |
|  | **Verbal reasoning** | |
|  | **Females** | **Males** |
| MBI syndrome |  |  |
| Model 1 | -0.300 (-0.586; -0.014) * | -0.292 (-0.560; -0.023)* |
| Model 2 | -0.301 (-0.586; -0.016) * | -0.292 (-0.560; -0.024)* |
| Model 3 | -0.299 (-0.585; -0.013) * | -0.290 (-0.559; -0.022)* |
| Emotional dysregulation |  |  |
| Model 1 | -0.177 (-0.301; -0.053)** | -0.165 (-0.366; 0.037) |
| Model 2 | -0.177 (-0.301; -0.053)** | -0.164 (-0.364; 0.037) |
| Model 3 | -0.174 (-0.298; -0.049)** | -0.164 (-0.366; 0.038) |
| Decreased motivation |  |  |
| Model 1 | -0.234 (-0.377; -0.091)** | -0.307 (-0.517; -0.097)** |
| Model 2 | -0.235 (-0.378; -0.092)** | -0.307 (-0.516; -0.098)** |
| Model 3 | -0.226 (-0.370; -0.083)** | -0.308 (-0.518; -0.098)** |
| Impulse dyscontrol |  |  |
| Model 1 | -0.138 (-0.265; -0.011)* | -0.182 (-0.375; 0.012) |
| Model 2 | -0.138 (-0.264; -0.011)* | -0.181 (-0.375; 0.012) |
| Model 3 | -0.136 (-0.263; -0.009)* | -0.176 (-0.371; 0.018) |
| Social inappropriateness |  |  |
| Model 1 | -0.220 (-0.413; -0.028)* | -0.282 (-0.552; -0.012)* |
| Model 2 | -0.222 (-0.414; -0.029)* | -0.281 (-0.551; -0.012)* |
| Model 3 | -0.219 (-0.412; -0.026)* | -0.266 (-0.537; 0.005) |
| Psychotic symptoms |  |  |
| Model 1 | -0.175 (-0.427; 0.076) | -0.448 (-0.882; -0.015)* |
| Model 2 | -0.174 (-0.425; 0.078) | -0.452 (-0.884; -0.019)* |
| Model 3 | -0.157 (-0.410; 0.096) | -0.449 (-0.882; -0.016)* |

*** p<0.001 ** p<0.01 * p<0.05; CI, confidence interval; MBI, mild behavioral impairment

Model 1: baseline age, practice effect

Model 2: baseline age, practice effect, employment status, ethnic origin, co-habitation status, education level

Model 3: baseline age, practice effect, employment status, ethnic origin, co-habitation status, education level, body-mass index, hypertension, history of heart disease, diabetes, hypercholesterolemia
